# Perceived constraints to healthy diets: Evidence from agrifood system assessments in rural South Asia

**DOI:** 10.1101/2025.01.06.25320037

**Authors:** Sharvari Patwardhan, Suman Chakrabarti, Esther M. Choo, Morgan Boncyk, Christine Blake, Sunny S. Kim, TAFSSA Collaborators, Samuel Scott

## Abstract

The healthfulness of diets in South Asia is limited by socio-economic and public infrastructure challenges. Perceptions about food such as availability, accessibility, desirability, and convenience can impact food choice and ultimately diets. However, there are limited tools to understand consumers’ perceptions of these factors and if perceptions relate to actual food intake. Using a novel tool administered across five rural districts in Bangladesh, India, and Nepal, we quantify the association between food perceptions and food intake. A Likert scale (agree, neutral, disagree) was used to capture respondents’ perceptions about seven food choice drivers (affordability, accessibility, desirability, convenience, food quality, food safety, availability) for a list of six common foods. For each food, principal Component Analysis (PCA) was used to identify latent “drivers”. The association between these and diets (using 24-hour dietary recall data) was estimated using multivariable regression analysis. There was considerable heterogeneity across countries with respect to the relative importance of food choice drivers and diet quality. There is a need to measure and understand individual food perceptions that drive food choice to help develop policies that promote healthier food choices.

## INTRODUCTION

Food choice behaviors or consumer behaviors are the activities which individuals engage with markets and how they purchase, use, and dispose of goods and services (International Food Policy Research Institute, 2024). Food choice behaviors are the outcomes of conscious and unconscious processes of food choice decision-making (Boncyk et al., 2023). Food choice is not limited to food consumption alone and can operate at the individual and household levels (Turner et al., 2018). It is deeply intertwined with expressions of identity, preferences, and socio-cultural values that ultimately shape dietary intake and health outcomes (Blake et al., 2021; Boncyk et al., 2023).

Perceptions of the external and personal food environment drive food choice and ultimately diets. For instance, the 2024 Global Food Policy Report identifies affordability, availability, accessibility, and desirability as key factors affecting food choice, yet there have been limited quantitative assessments of how individual perceptions of these domains relate to diet quality (International Food Policy Research Institute, 2024). Understanding how individual perceptions about certain food environment attributes relate to consumption behavior and diet quality is especially important in the South Asian context where diets are limited by poverty, high prices relative to income, availability and access issues, among others (FAO et al., 2024). Further, with increasing consumption of unhealthy foods across South Asia (Gillespie et al., 2019), understanding the drivers of food choice at the individual level is essential for developing programs and policies to improve diets (Karanja et al., 2022).

Drivers of food choice can be measured from the market side and the household side (Turner et al., 2018). There is a larger body of work on the external food environment (Constantinides et al., 2021), however, less is known about food choice drivers from the individual perspective. For instance, availability refers to the level of presence of a food source or the product itself (Charreire et al., 2010; Lake, 2018). Availability can be measured by the varieties and quantities of different types of food sold in community markets. However, even if a food is available near where someone lives, they may not know about it, or it may sell out quickly, or only be offered at certain times of year. There may not be an exact match when measuring availability objectively in a market setting versus measuring availability from an individual’s perspective. In other words, individuals may have access to more relevant information that influences their food choice.

We implemented a novel tool for measuring individual perceptions of food choice drivers among adults and adolescents to quantitatively examine the association between food choice drivers and diet quality and food consumption across five rural districts in three countries in South Asia.

## METHODS

### Description of survey

District-representative individual-level quantitative data were collected in February-May 2023 from Nalanda district in Bihar state (India); Surkhet district in Lumbini Province and Banke district in Karnali Province (Nepal); and Rangpur district in the Rangpur division and Rajshahi district in the Rajshahi Division (Bangladesh) under the Consultative Group for International Agricultural Research (CGIAR) initiative ‘Transforming Agrifood Systems in South Asia’ (TAFSSA) (Gupta et al., 2022). These locations were selected as they face common challenges related to poverty, climate, malnutrition and social inclusion, and were known sites with previous CGIAR research investments. The surveys were designed to be representative at the district level. The sampling approach and sample size determination followed the example of India’s equivalent of the Demographic and Health Surveys, the National Family Household Survey (NFHS), a population-based survey representative at the district level most recently conducted in 2019-2021. We exceeded the number of households per district in NFHS and used a sample size of 1000 households per district for our survey. In Nepal, relative to Bangladesh and India, districts were much less populated, thus we included 500 households per district in Nepal.

The primary sampling unit (PSU) was villages (in Bangladesh and India) and wards (in Nepal). PSUs were selected from the most recent National Census datasets from each country, using a Probability Proportional to Size (PPS) sampling strategy. In the first stage of sampling, villages or wards were selected at random, with a probability proportional to the number of households that reside in the PSU. In the second stage, an equal number of households were selected randomly from each PSU, roughly 20 households per PSU. The sampling frame included all rural households that included an adolescent member. The rationale for only including households with adolescents was to better understand adolescents’ roles in agrifood systems which include the activities and actors involved in getting food from farm to plate.

At the household level, one adult female aged 20+ years, one adult male aged 20+ years, and one adolescent respondent aged 10-19 years regardless of sex were invited as respondents. In case more than one adolescent lived in the household, the oldest adolescent was selected as the respondent. The adult female respondent was identified as the female household member primarily responsible for managing the household. The adult male respondent was identified as the male household member primarily responsible for agricultural activities. If one person was responsible for both activities, the same individual responded to both the female and male questionnaires.

Three survey firms supported data collection: Data Analysis and Technical Assistance in Bangladesh, Kabil Professional Services in India, and Institute of Integrated studies in Nepal. After explaining the survey’s objectives and procedures, enumerators obtained verbal informed consent from respondents. Participation in the survey was voluntary and participants were free to end interviews at any time by informing survey staff. The protocol, informed consent forms, and study questionnaires were approved by the Institutional Review Boards (IRB) of the International Food Policy Research Institute in Washington, DC, USA; the Institute of Health Economics, University of Dhaka, Bangladesh; Centre for Media Studies, Delhi, India; and the National Health Research Council, Kathmandu, Nepal.

### Data collection procedure for individual perceptions of food choice drivers

For each respondent, a novel tool adapted from Choo’s food environment rapid assessment (FERA) tool (Choo, 2023) was implemented to collect data on individual perceptions of food choice drivers. Respondents were requested to agree/disagree/neither to statements (**Table 1**) related to seven individual-level food choice drivers (affordability, accessibility, desirability, convenience, food quality, food safety, and availability) for a pre-determined list of six “sentinel” foods/food groups common across the study countries (lentils, eggs, green leafy vegetables, banana, biscuits, and fried food). The seven statements were read for a single food and then repeated for each food in sequence. The selection of sentinel foods/food groups was based on scoping visits to field sites to understand commonly consumed foods across different food groups.

**Table 1:**
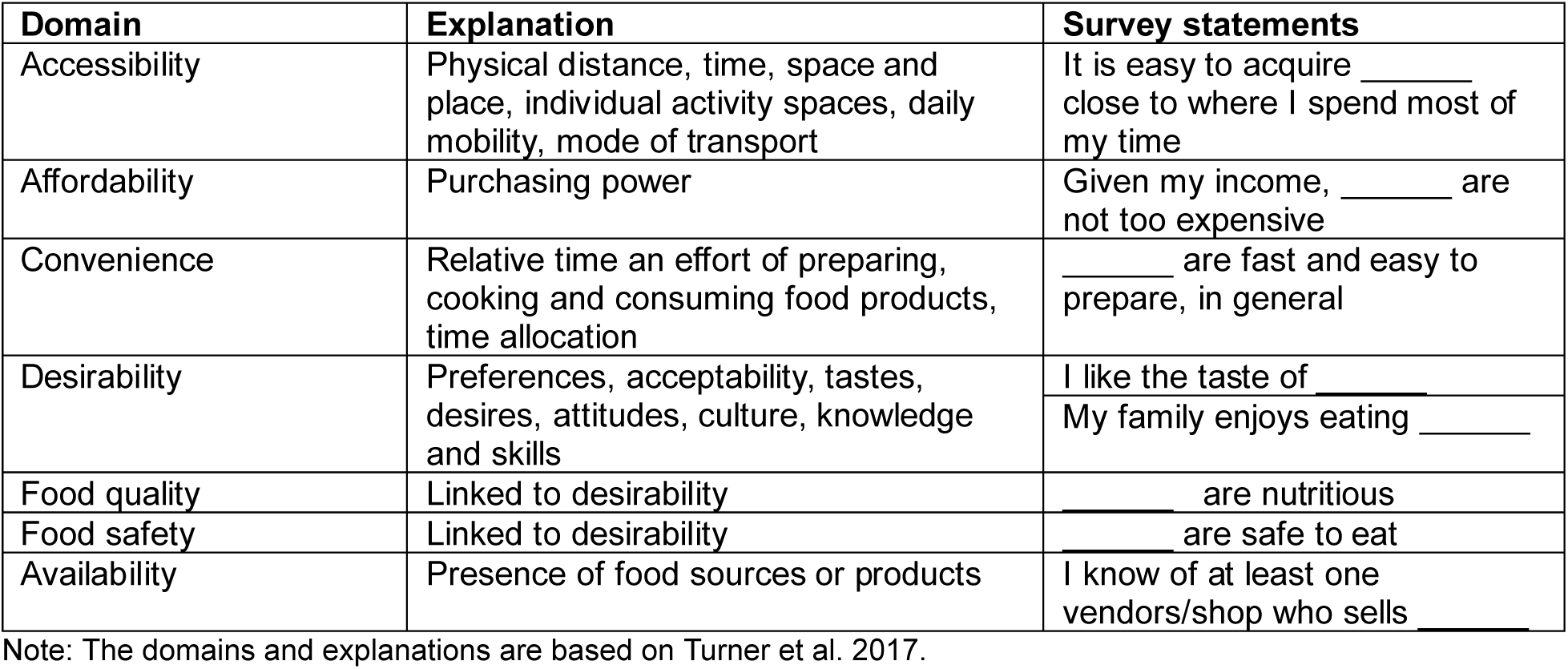
Mapping survey statements to the domains in the framework proposed by Turner et al.^4^.

The original food environment rapid assessment (FERA) tool (Choo, 2023) consisted of 14 statements related to ten food choice drivers on the external (availability, stability, product properties, vendor properties, marketing) and internal (accessibility, affordability, convenience, desirability – including food quality and food safety, social forces) domains, which were developed based on Turner’s Framework (Turner et al., 2018). Previously the FERA was only used with two food items, but we were interested in understanding perceptions of a wider range of foods. To keep the length of the tool reasonable in a household survey with many other modules, we reduced the number of statements from 14 to 8, primarily by dropping statements that mapped to some external drivers domain, which were measured objectively elsewhere in the survey. During pre-testing, some statements from the FERA tool were reworded in local languages to improve respondent comprehension.

### Dietary assessment

The Global Diet Quality Score (GDQS) application (Moursi et al., 2021) was used to collect 24-hour ingredient-level dietary recall data, used to produce the GDQS metrics. The GDQS metrics have been validated to predict nutrient inadequacy and non-communicable disease (NCD) risk outcomes globally. The metric considers the level of consumption of 25 food groups: 16 healthy food groups, 2 food groups that are unhealthy when consumed in excess, and 7 unhealthy food groups. Points are assigned based on the level of consumption, with more points for higher consumption of healthy foods and lower consumption of unhealthy foods. The GDQS plus score is the sum of points received from consuming healthy food groups and has a possible range of 0-32 points. A higher GDQS plus score indicates higher consumption of healthy foods. The GDQS minus score is the sum of points received from consuming unhealthy food groups including the two food groups that are unhealthy when consumed in excessive amounts and has a possible range of 0-17 points. A higher GDQS minus score indicates lower consumption of unhealthy foods. In addition to the diet quality scores, we examined consumption of the six individual sentinel foods that the perception questions were based on. Data on consumption of the six sentinel foods/food groups in the past 24 hours were extracted from the GDQS application.

A food frequency questionnaire (FFQ) was administered to collect 7-day dietary recall data. Respondents were asked how often they consumed each sentinel food prepared at home or outside, in the past 7 days. Response options ranged from never (0 times) to 6 or more times per day (truncated to 42 times per week).

### Statistical analysis

#### Principal component analysis to identify latent constructs of food perceptions

The individual FERA statements were mapped a-priori to the domains of drivers of food choice (**Table 1**). The responses to statements were assigned a score (agree=1, disagree=0, neither agree nor disagree=0.5). We summed the scores for the 4 healthy foods (lentils, eggs, green leafy vegetables, banana) to produce a ‘healthy score’ and did the same for the 2 unhealthy foods (biscuits, fried food) to produce an ‘unhealthy score’. The healthy and unhealthy scores were standardized by dividing by the number of foods/food groups in that score (the healthy score was divided by 4 and the unhealthy score by 2). The score of 0 represented the minimum level of the domain, and 1 represented the maximum.

Due to the presence of collinearity between responses to the individual FERA statements, a Principal Component Analysis (PCA) approach was used to understand which combinations of the responses maximally explain the variance in the sample and capture the theoretical latent domains of food perceptions. The components derived from the PCA were an a-posteriori estimation of the a-priori mapping of the responses as displayed in **Tables S2-S3**. In other words, the PCA allowed us to statistically test whether the responses resulted in consistent estimation of domains listed in the FERA tool (Choo, 2023).

We conducted eight different PCA models. The first two PCA models were conducted using the responses to statements on 1) healthy foods (lentils, eggs, green leafy vegetables, banana) and 2) unhealthy foods (biscuits, fried foods). The principal components (PCs) obtained were scaled, ranging from 0-1, with 0 representing the lowest level of a food choice driver domain and 1 representing the highest. For sensitivity checks, to understand if perceptions of individual sentinel foods were related to consumption of those individual foods, PCAs were also conducted for each of the six sentinel foods separately. To understand the latent food choice driver derived from each PC, we considered factor loadings >0.3 (Tabachnick & Fidell, 2013). However, all loadings were considered in the factor score used in the regression analyses.

#### Regression analyses

The PCA-derived latent food choice drivers were examined against the outcome variables – GDQS plus, GDQS minus, and consumption of the six individual sentinel foods in the past 24 hours. The outcomes were standardized as country specific z-scores to aid interpretation of the magnitude of the associations for the local country context. The association between the food choice drivers’ points derived from the PCA and GDQS was estimated using Ordinary Least Squares (OLS) regression models. These models adjusted for gender, age, wealth (the index was pooled for all countries), years of education, household size, food insecurity, and included district fixed effects.

The wealth index was calculated using asset-based information including ownership of durable goods, housing characteristics, access to clean cooking fuel, and access to improved toilet facilities among others. This information was pooled for households from all three countries. Food insecurity was measured using the food insecurity experiences scale with 8 items and a 30-day recall, using a cutoff of ≥1 to indicate food insecure. District fixed effects accounted for all unobserved time-invariant confounders at the district level such as geography and climate among others. Standard error estimates were clustered at the household level to account for outcome correlations within households. OLS regression coefficients are interpreted as the difference in the standard deviations of the outcome comparing the lowest level of the explanatory variable to the highest level. We use logistic models with the same specification to examine the association between food-specific drivers and consumption of each sentinel food (yes or no) in the past 24 hours.

All results are reported by country (not district) for ease of interpretation and reporting. Further, we first ran models separately by respondent (adult male, adult female, adolescent male and female) but did not find any meaningful differences, thus we report findings for all respondent types combined. All analyses were performed in Stata version 18.

#### Sensitivity analysis

Even if a respondent did not consume a sentinel food in the past 24 hours, they may have consumed that food in the past 7 days. Examining frequency rather than any consumption in the past 7 days also provided more variation in the outcome. We used the number of times a sentinel food was consumed in the past 7 days (ranging from 0-42) as an outcome to account for habitual eating patterns rather than recent (past 24 hours) consumption. We conducted this supplementary analysis to check the sensitivity of the food choice drivers to changing the outcome from 24-hour dietary recall to 7-day dietary recall.

## RESULTS

### Respondent characteristics, diet, and perceptions of food choice drivers

The total sample size across the locations was 9,702 respondents (**Table 2**). On average, respondents were 30-32 years old with 5-6 years of education. A higher percentage of households were female-headed in Nepal (39%) compared to India (21%) and Bangladesh (9%). While food insecurity was common in all three countries, India had the highest percentage of food insecure households (72%). A higher percentage of respondents from Bangladesh (24%) were in the lowest wealth quintile compared to India (18%) and Nepal (15%). When examining the average number of times of sentinel foods consumption in the past week, lentils were more frequently consumed across countries. Biscuits and fried foods were more frequently consumed in Bangladesh compared to India and Nepal.

**Table 2:**
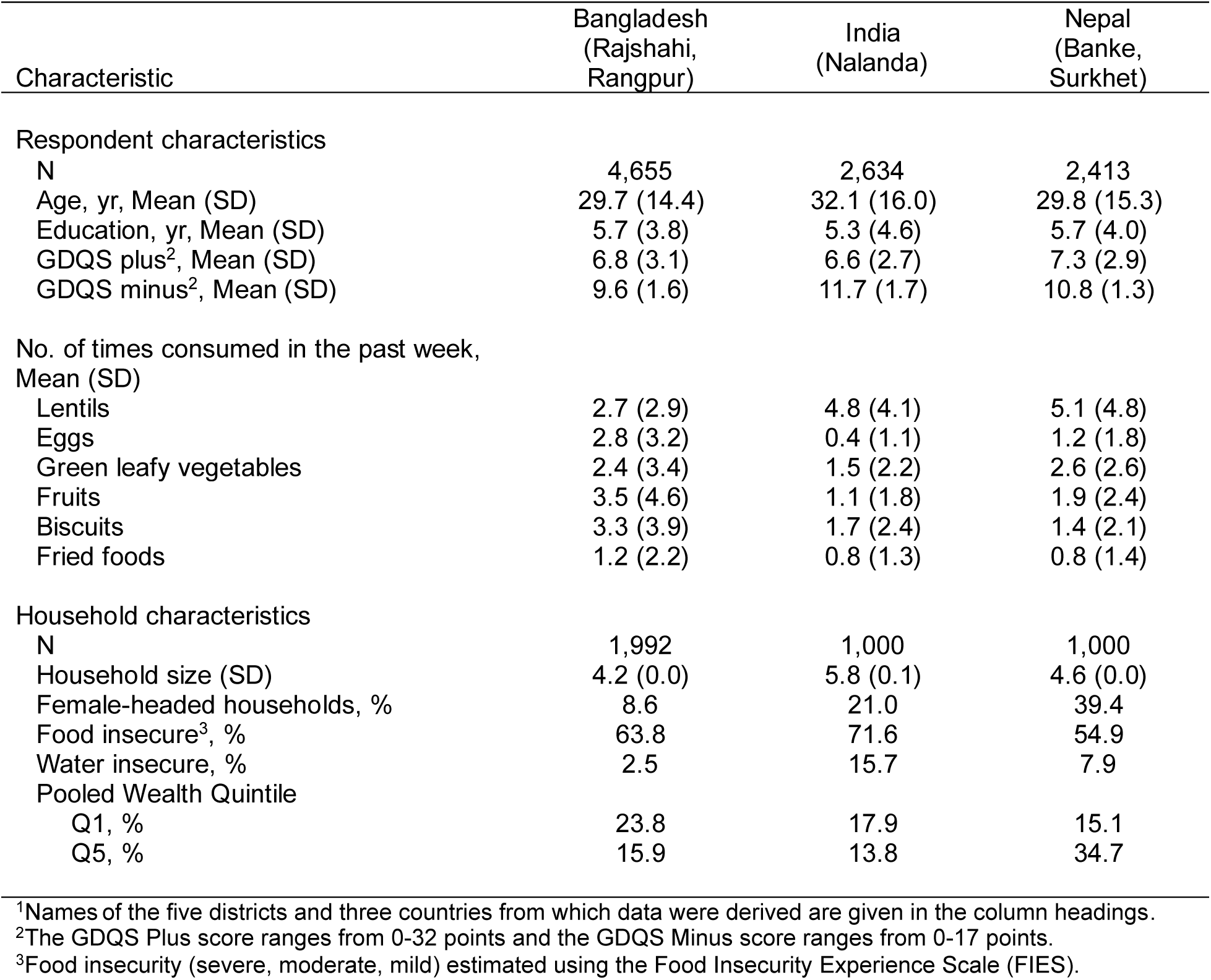
Sample characteristics in Bangladesh, India, and Nepal^1^.

Respondents in Nepal consumed more healthy foods (GDQS+: 7.3) compared to respondents in Bangladesh (GDQS+: 6.8) and India (GDQS+: 6.6) (**Table 2**). Respondents in India consumed less unhealthy foods (GDQS-: 11.7) compared to respondents in Nepal (GDQS-: 10.8) and Bangladesh (GDQS-: 9.6). Lentils were the most consumed sentinel food and bananas were the least consumed, with consumption patterns varying by country (**Figure 1**).

**Figure 1:**
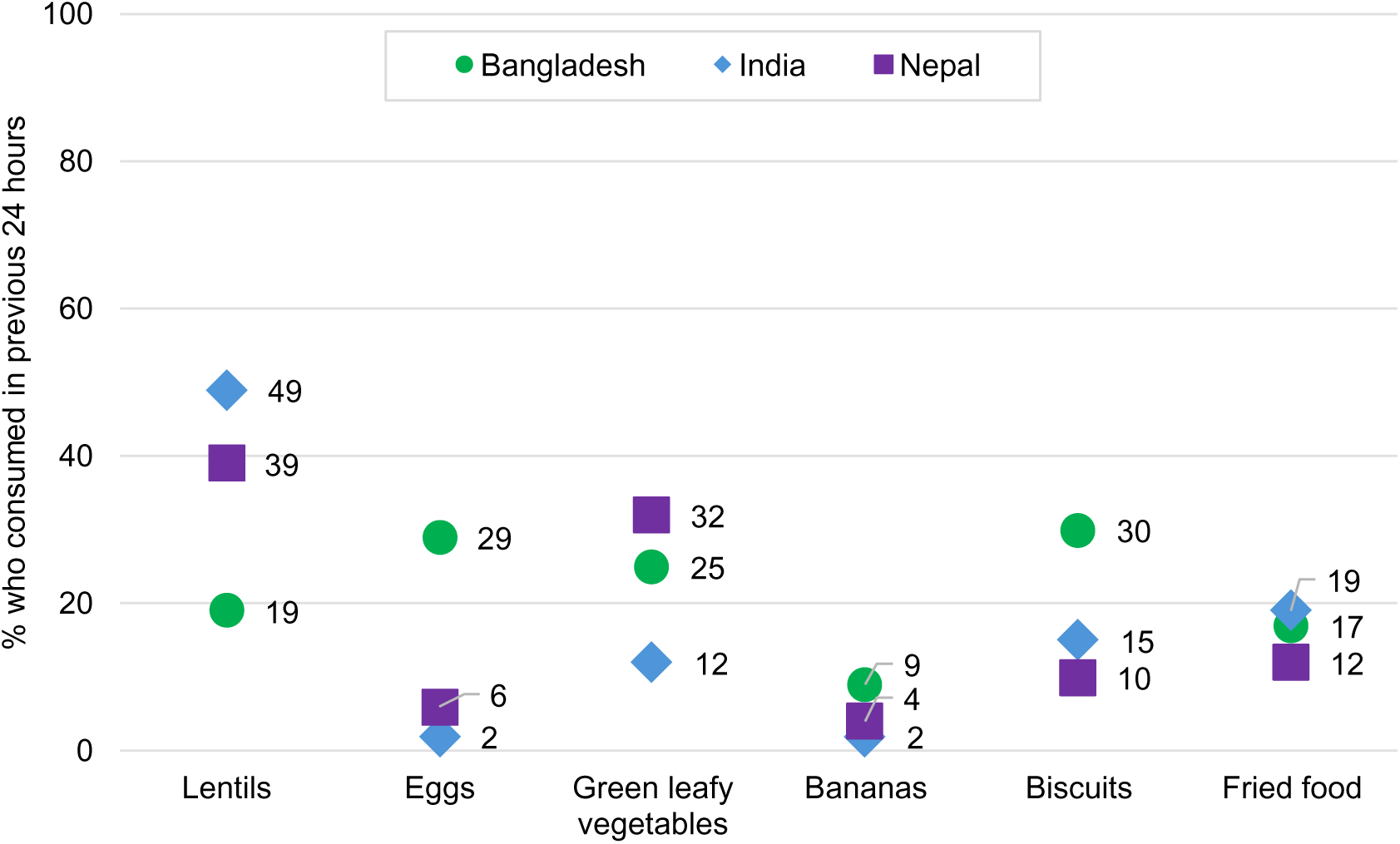
Percentage of respondents who consumed different types of foods in the past 24 hours.

Most respondents felt that healthy foods were desirable, nutritious, and safe to eat and that unhealthy foods were easily available (**Table S1**). Only 16% (Bangladesh) to 43% (India) agreed to the statement that fried foods were nutritious and 17% (Bangladesh) to 37% (India) agreed that fried foods were safe to eat. In Bangladesh, 72%-93% (depending on the type of food) of respondents perceived that foods were affordable, compared to 17%-65% in India and Nepal.

### PCA Results

The estimates derived from the PCA conducted using the indicators for the healthy sentinel foods (lentils, eggs, green leafy vegetables, and banana) are displayed in **Table S2**. PC1 loaded on three food choice driver statements – like the taste of (factor loading: 0.69), is nutritious (0.35), and family enjoys eating (0.61) healthy foods – which were theoretically mapped to and correspondingly labelled “desirability”. PC2 was labelled “affordability and accessibility” as the components loaded on not expensive (0.81) and easy to acquire (0.47) healthy foods. The remaining Principal Components were similarly labelled “food safety and quality” (PC3), “availability and accessibility” (PC4), and “convenience” (PC5). A similar exercise was conducted to label the domains derived from the PCA conducted with unhealthy foods (**Table S3**). The PCA model with the unhealthy foods resulted in components that load on the statements which were similar to the PCA model with healthy foods. Overall, PCA results (across all PCA models) concurred with the theoretical DFC domains (**Tables S2-S9**).

### Association between constructs of food perceptions and diet quality

Perceived affordability, availability, accessibility, and desirability of healthy foods predicted higher intake of these foods (**Figure 2**). After adjustment for covariates, in India, every unit increase in affordability-accessibility, convenience, and desirability predicted a higher GDQS plus z-score by 0.33 (95% CI 0.164, 0.509), 0.25 (95% CI 0.092, 0.408), and 0.12 (95% CI 0.003, 0.234) points respectively (p<0.05). Because GDQS plus is normally distributed, a positive change in z-scores of 0.7 points (summing 0.33, 0.25, and 0.12) would account for 30% of the variation under the bell curve. Similarly, in Bangladesh, every unit increase in perceived affordability-accessibility, and availability-accessibility was associated with a higher GDQS plus z-score by 0.19 (95% CI 0.028, 0.355) and 0.12 (95% CI -0.004, 0.246) points respectively (p<0.05). However, in Nepal, only higher perceived desirability significantly predicted a higher GDQS plus z-score by 0.41 (95% CI 0.190, 0.628) points (p<0.05).

**Figure 2:**
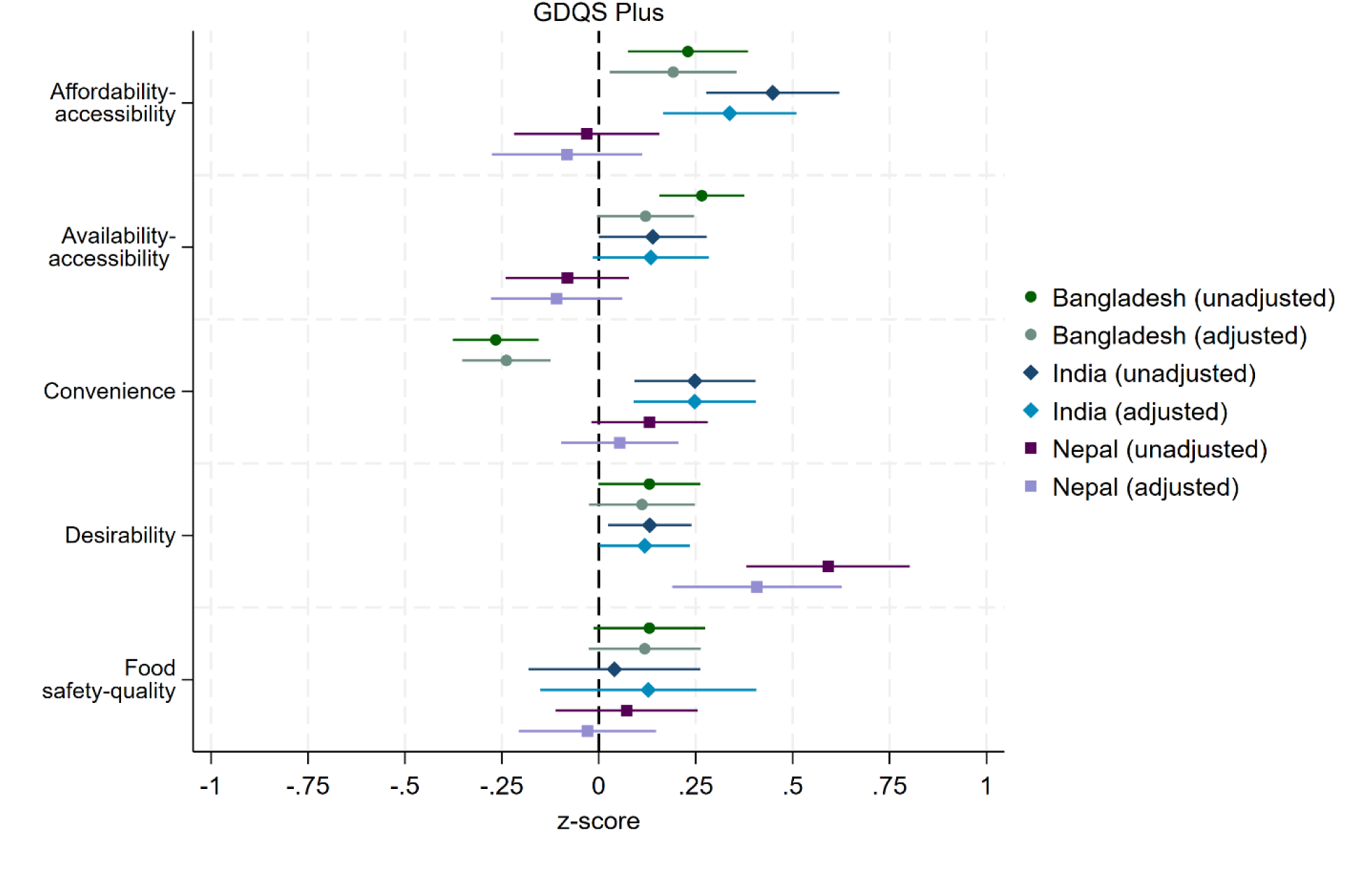
Unadjusted and adjusted OLS estimates of GDQS Plus z-score by the drivers of healthy foods^1^, by country. Note: The GDQS Plus score ranges from 0-32 points, summed across 16 healthy food groups; higher scores indicate higher consumption of healthy foods. The adjusted model includes controls for gender, age, years of education, household size, food insecurity, wealth, and district fixed effects. Standard errors have been clustered at the household level. ^1^See results on the drivers of healthy foods in **Table S3**.

Availability, accessibility, and desirability of unhealthy foods were significant predictors of higher unhealthy food consumption (**Figure 3**). In Bangladesh, every unit increase in perceived availability-accessibility were associated with a lower GDQS z-minus score by -0.18 points (95% CI -0.284,-0.066; p<0.05). In India, every unit increase in perceived availability-accessibility and desirability predicted a lower GDQS minus z-score by -0.28 (95% CI -0.405, -0.146) and -0.26 (95% CI -0.412, -0.110) points respectively (p<0.05). Similarly, in Nepal, every unit increase in perceived desirability significantly predicted a lower GDQS minus z-score by -0.34 points (95% CI -0.515,-0.156; p<0.05).

**Figure 3:**
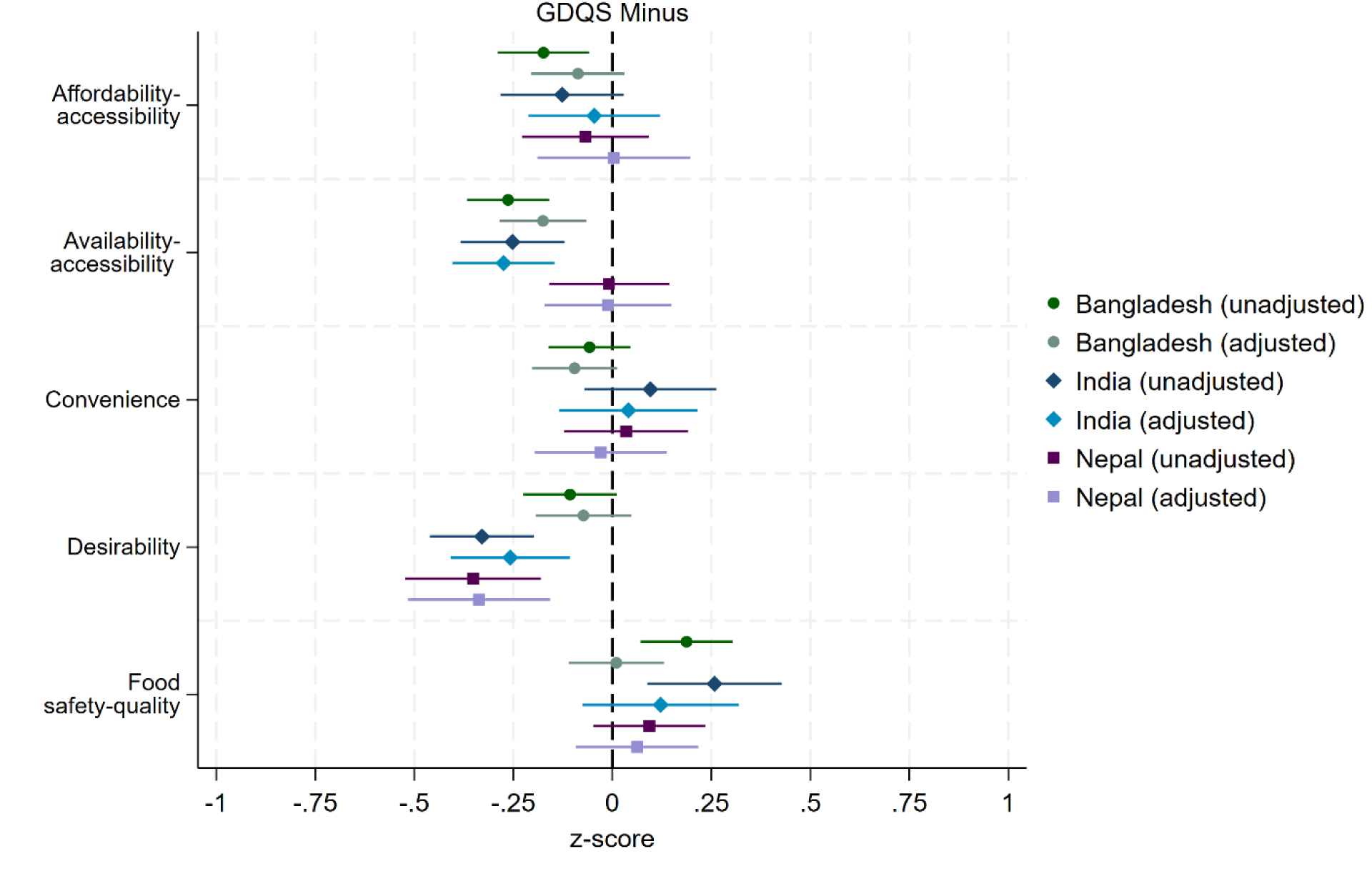
Unadjusted and adjusted OLS estimates of GDQS Minus z-score by the drivers of unhealthy foods^1^, by country. Note: The GDQS minus score ranges from 0-17 points, summed across 7 unhealthy food groups and 2 food groups that are unhealthy when consumed in excessive amounts; higher scores indicate lower consumption of unhealthy foods. The adjusted model includes controls for gender, age, years of education, household size, food insecurity, wealth, and district fixed effects. Standard errors have been clustered at the household level. ^1^See results on the drivers of unhealthy foods in **Table S4.**

In the food-specific analysis, a higher consumption of biscuits and fried foods was associated with higher perceived accessibility, affordability, convenience, desirability, and food safety-quality, across countries (**Table 3**). For healthy foods consumption, higher perceived desirability predicted a higher consumption of lentils (India), eggs (India and Nepal), green leafy vegetables (Bangladesh and Nepal), and bananas (Bangladesh). While greater perceived convenience and desirability were associated with higher consumption of lentils, eggs, green leafy vegetables, and bananas, more food choice driver perceptions were associated with consumption of both biscuits and fried food.

**Table 3:**
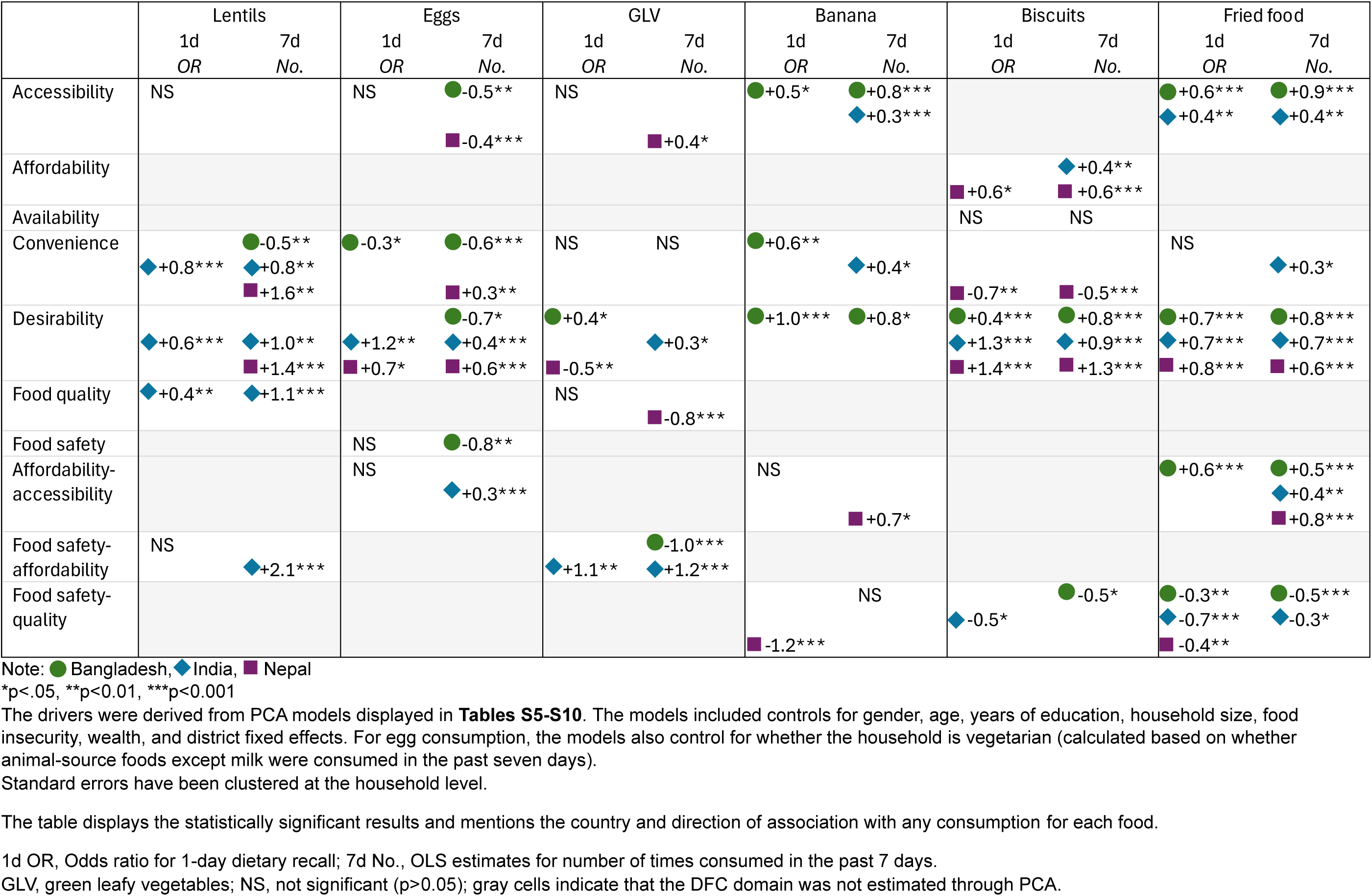
Summary of logit and OLS estimates for 1-day versus 7-days sentinel foods consumption respectively by drivers.

### Results of the sensitivity analysis

We observe some differences in the estimates for any consumption in the past 24 hours compared with frequency of consumption in the past 7 days (**Table 3**). For unhealthy foods, the results were similar for 7-day versus 1-day consumption. However, for lentils, eggs, and green leafy vegetables, frequency of consumption in the past 7 days is associated with more factors including food safety, affordability-accessibility, and food safety-affordability. This shows that the perceptions of food choice drivers have varied association with differing periods of consumption. When examining consumption over a longer period of time, more food choice driver perceptions were associated with consumption of healthy and unhealthy foods.

## DISCUSSION

This study implemented a novel tool to quantitatively examine subjective perceptions about food choice drivers. We found that the tool effectively captured food choice driver domains that successfully predicted diet quality and consumption. The analysis underscored the importance of desirability perceptions in determining food choice. Desirability was the most commonly occurring factor associated with diet quality and consumption across countries and food groups. The analysis also showed that a higher number of factors (such as affordability and food safety) predicted unhealthy food intake. This suggested that unhealthy foods are responsive to changes in a larger set of food choice driver perceptions than healthy foods. Further, there was substantial heterogeneity in the relative importance of these factors across countries, demonstrating the need for context-specific research. This study highlights the utility of applying tools that measure the perceptions of food choice drivers and their implementation across different country contexts.

The strengths of this study lie in the development and use of a time-saving tool that is aligned with Turner’s food environment framework (Turner et al., 2018) and in estimating subjective perceptions of the personal food environment domains. The tool can further be easily adapted to varied contexts by appropriately translating the questions and using a tailored list of sentinel foods. Secondly, we applied rigorous statistical methods to extract latent food choice driver constructs and created statistical indicators to quantitatively examine their relationship with diet quality and food consumption. Moreover, we used a robust set of covariates to account for plausible confounding that may have otherwise provided biased estimates of the association between food choice drivers and diets. Lastly, the data used from diverse South Asian contexts had high external validity and the extracted food choice drivers were quite consistent across these contexts.

The study is limited by the subjectivity in the list of sentinel foods included, the indicators used in the PCA, and the interpretation of the PCA domains. For example, the choice of conducting separate PCAs for healthy and unhealthy foods may have influenced results. Further, while the analysis predicted strong associations, causal relationships cannot be inferred from the cross-sectional data that were used. The results were also dependent on the choice of outcome. We found differences in significance and magnitude of drivers when looking at daily versus weekly dietary recall. Lastly, respondents across countries may have interpreted questions from the adapted FERA tool differently than intended. For example, questions about food safety may have been interpreted as long term health implications of consuming a particular food rather than hygiene-related safety. Responses from Bangladesh were also observed to be conforming more towards a social desirability bias. Spuriously high agreement rates to some questions may have biased results towards the null.

As expected, we found that association in the 7-day recall were more pronounced than the 24-hour recall because consumers are likely to eat a higher variety of foods through the week. In rural contexts, where access to refrigeration and other means of food preservation are limited, households consume a repetitive set of foods throughout the day such as rice, lentils, seasonal vegetables (Scott et al., 2023). Our findings suggest that South Asian rural households may be changing healthy foods across days of the week rather than within a single day making it challenging to choose between methods that rely on 24-hour versus 7-day dietary recall. However, for unhealthy foods, particularly biscuits which are consumed daily with tea, we detected significant associations in both sets of recall.

There was considerable heterogeneity across countries with respect to the relative importance of food choice drivers and diet quality obtained from the GDQS indicators. In Bangladesh, individuals with greater perception of affordability and availability ate more healthy foods but those led by convenience did not. In India, healthy foods that were perceived to be affordable and convenient were consumed the most, whereas unhealthy foods were consumed if they were perceived to be available and desirable. Food choices of individuals in Nepal were largely determined by perceptions of desirability for healthy and unhealthy foods.

In our study, adults and adolescents, regardless of sex, appeared to have similar associations among drivers and diets implying that there may be some homogeneity in consumption within these households. For example, the strong associations of desirability with the consumption of unhealthy foods suggests that rural South Asian populations are at risk of susceptibility to advertising from traditional and modern media. On the other hand, media could also be leveraged to promote the desirability for healthy foods in rural populations. Public health platforms and community events could also be leveraged to deliver nutrition-focused behavior change communication (Kachwaha et al., 2024).

The study provides important insights into factors that influence diets. While dealing with challenges such as accessibility and affordability constraints, rural South Asian diets appear to be influenced by desirability and convenience as well. The interaction and interconnectedness of these factors must be closely examined to probe into context-specific drivers of food choice. In this way, understanding perceptions of food that drive food choice can aid interventions for improving household diets.

## DATA AVAILABILITY

The datasets analyzed during this study are publicly available and can be accessed at https://www.cgiar.org/news-events/news/open-access-agrifood-system-data-from-4000-households-across-bangladesh-india-and-nepal/

---

**Table S1:** Percentage of respondents that agreed to the following statements for different foods.

|  | ACCESSIBILITY | AFFORDABILITY | CONVENIENCE | DESIRABILITY | DESIRABILITY | FOOD QUALITY | FOOD SAFETY | AVAILABILITY |
| --- | --- | --- | --- | --- | --- | --- | --- | --- |
|  | It is easy to acquire _____ close to where I spend most of my time | Given my income, _____ are not too expensive. | _____ are fast and easy to prepare, in general | I like the taste of _____ | My _____ family enjoys eating _____ | _____ are nutritious | _____ are safe to eat | I know of at least one vendors/shop who sells _____ |
| Rajshahi, Rangpur (Bangladesh) N=4,655 |  |  |  |  |  |  |  |  |
| Lentils, % | 86 | 76 | 88 | 91 | 95 | 98 | 98 | 85 |
| Eggs, % | 91 | 75 | 94 | 96 | 98 | 99 | 98 | 90 |
| GLV, % | 89 | 93 | 91 | 94 | 97 | 99 | 98 | 73 |
| Banana, % | 80 | 76 | 75 | 94 | 97 | 99 | 97 | 74 |
| Biscuits, % | 91 | 83 | 76 | 85 | 88 | 71 | 75 | 93 |
| Fried food, % | 70 | 72 | 64 | 67 | 66 | 16 | 17 | 75 |
| Nalanda (India) N=2,634 |  |  |  |  |  |  |  |  |
| Lentils, % | 85 | 20 | 86 | 92 | 92 | 97 | 98 | 93 |
| Eggs, % | 77 | 31 | 76 | 78 | 76 | 83 | 67 | 87 |
| GLV, % | 74 | 45 | 87 | 93 | 94 | 96 | 97 | 87 |
| Banana, % | 45 | 45 | 38 | 91 | 91 | 94 | 88 | 77 |
| Biscuits, % | 91 | 60 | 31 | 87 | 81 | 64 | 63 | 97 |
| Fried food, % | 61 | 32 | 58 | 81 | 78 | 43 | 37 | 86 |
| Banke, Surkhet (Nepal) N=2,413 |  |  |  |  |  |  |  |  |
| Lentils, % | 84 | 17 | 79 | 90 | 93 | 95 | 97 | 95 |
| Eggs, % | 88 | 29 | 91 | 88 | 90 | 93 | 88 | 96 |
| GLV, % | 80 | 51 | 92 | 92 | 94 | 98 | 99 | 87 |
| Banana, % | 71 | 31 | 89 | 95 | 96 | 96 | 95 | 94 |
| Biscuits, % | 96 | 65 | 91 | 67 | 72 | 36 | 36 | 99 |
| Fried food, % | 68 | 29 | 66 | 76 | 80 | 35 | 30 | 93 |

Note: The tool has been adapted from Choo (2023)<sup>6</sup>. GLV, green leafy vegetables.

**Table S2:** PCA estimates using indicators for four healthy foods (lentils, eggs, green leafy vegetables, and banana)

| Component | PC1 | PC2 | PC3 | PC4 | PC5 |
| --- | --- | --- | --- | --- | --- |
| Domain | Desirability | Affordability-accessibility | Food safety and quality | Availability-accessibility | Convenience |
| Eigenvalue | 2.45 | 1.18 | 1.14 | 0.8 | 0.67 |
| Variation explained, % | 20 | 15 | 15 | 14 | 14 |
| Indicator names |  |  |  |  |  |
| Not expensive healthy | 0.0548 | **0.8054** | 0.1066 | -0.1304 | -0.0843 |
| Know vendor healthy | -0.0052 | -0.0781 | 0.0149 | **0.9022** | -0.0841 |
| Like the taste of healthy | **0.6924** | 0.0718 | -0.1479 | -0.0031 | -0.0529 |
| Easy to acquire healthy | 0.0164 | **0.4687** | -0.062 | **0.403** | 0.2956 |
| Easy to prepare healthy | -0.0143 | -0.0343 | -0.0065 | -0.0635 | **0.9386** |
| Safe to eat healthy | -0.1256 | 0.1427 | **0.8271** | 0.0001 | -0.0407 |
| Is nutritious healthy | **0.3513** | **-0.3118** | **0.5272** | 0.0472 | 0.1144 |
| Family enjoys eating healthy | **0.6148** | 0.0407 | 0.0264 | -0.0165 | 0.0067 |

Note: The significant domains are based on a cutoff of 0.3 for the factor loadings.

**Table S3:**
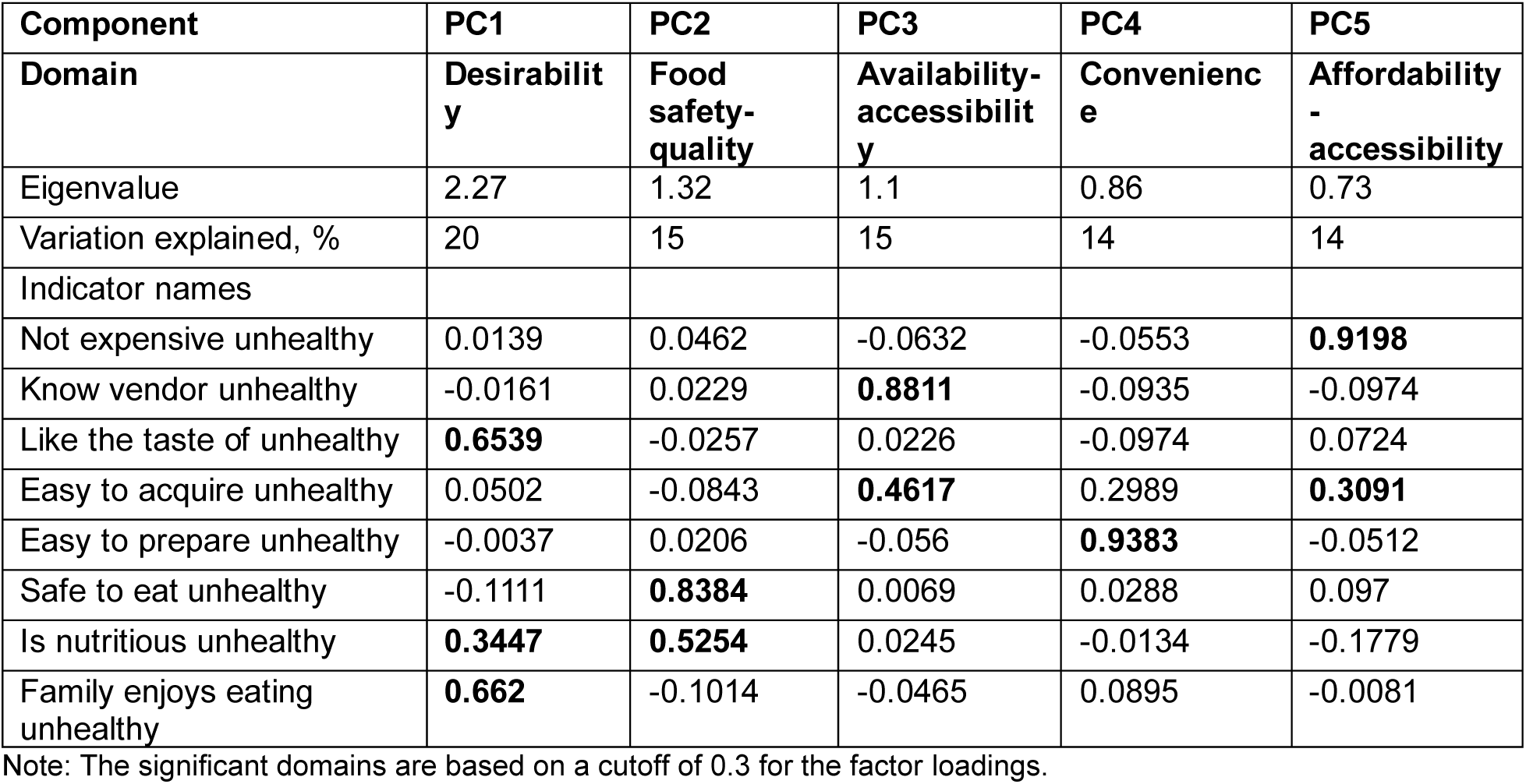
PCA estimates using indicators for two unhealthy foods (biscuits and fried food)

| Component | PC1 | PC2 | PC3 | PC4 | PC5 |
| --- | --- | --- | --- | --- | --- |
| Domain | Desirability | Food safety-quality | Availability-accessibility | Convenience | Affordability-accessibility |
| Eigenvalue | 2.27 | 1.32 | 1.1 | 0.86 | 0.73 |
| Variation explained, % | 20 | 15 | 15 | 14 | 14 |
| Indicator names |  |  |  |  |  |
| Not expensive unhealthy | 0.0139 | 0.0462 | -0.0632 | -0.0553 | **0.9198** |
| Know vendor unhealthy | -0.0161 | 0.0229 | **0.8811** | -0.0935 | -0.0974 |
| Like the taste of unhealthy | **0.6539** | -0.0257 | 0.0226 | -0.0974 | 0.0724 |
| Easy to acquire unhealthy | 0.0502 | -0.0843 | **0.4617** | 0.2989 | **0.3091** |
| Easy to prepare unhealthy | -0.0037 | 0.0206 | -0.056 | **0.9383** | -0.0512 |
| Safe to eat unhealthy | -0.1111 | **0.8384** | 0.0069 | 0.0288 | 0.097 |
| Is nutritious unhealthy | **0.3447** | **0.5254** | 0.0245 | -0.0134 | -0.1779 |
| Family enjoys eating unhealthy | **0.662** | -0.1014 | -0.0465 | 0.0895 | -0.0081 |

Note: The significant domains are based on a cutoff of 0.3 for the factor loadings.

**Table S4:** PCA estimates for indicators for “Lentils”.

| Lentils |  |  |  |  |  |
| --- | --- | --- | --- | --- | --- |
| Component | PC1 | PC2 | PC3 | PC4 | PC5 |
| Domain | Food safety-affordability | Desirability | Accessibility | Convenience | Food quality |
| Eigenvalue | 1.58 | 1.28 | 1.17 | 0.96 | 0.87 |
| Variation explained, % | 16 | 16 | 15 | 14 | 13 |
| Indicator names |  |  |  |  |  |
| Lentils_Not expensive | **0.6286** | -0.0498 | -0.1079 | **0.3199** | -0.0315 |
| Lentils_Know vendor | -0.0369 | -0.0109 | **0.7999** | -0.1507 | 0.0318 |
| Lentils_Like the taste of | -0.0262 | **0.6751** | -0.0083 | -0.0207 | 0.0970 |
| Lentils_Easy to acquire | 0.0888 | 0.0136 | **0.5861** | **0.3479** | -0.0571 |
| Lentils_Easy to prepare | -0.0707 | 0.0118 | -0.0434 | **0.8350** | 0.0358 |
| Lentils_Family enjoys eating | 0.0291 | **0.7349** | -0.0006 | 0.0299 | -0.0858 |
| Lentils_Safe to eat | **0.7675** | 0.0350 | 0.0547 | -0.2346 | 0.0338 |
| Lentils_Is nutritious | 0.0077 | -0.0045 | 0.0051 | 0.0176 | **0.9877** |

Note: The significant domains are based on a cutoff of 0.3 for the factor loadings.

**Table S5:** PCA estimates for indicators for “Eggs”.

| Eggs |  |  |  |  |  |
| --- | --- | --- | --- | --- | --- |
| Component | PC1 | PC2 | PC3 | PC4 | PC5 |
| Domain | Desirability | Accessibilit<br>y | Convenienc<br>e | Affordability<br>-<br>accessibility | Food<br>safety |
| Eigenvalue | 2.58 | 1.12 | 1.02 | 0.8 | 0.7 |
| Variation explained, % | 23 | 15 | 14 | 13 | 13 |
| Indicator names |  |  |  |  |  |
| Eggs_Not expensive | 0.0009 | -0.0526 | -0.0442 | **0.9393** | 0.0561 |
| Eggs_Know vendor | -0.0254 | **0.8438** | -0.1272 | -0.1248 | 0.0957 |
| Eggs_Like the taste of | **0.6539** | -0.0178 | -0.0786 | 0.0442 | -0.0758 |
| Eggs_Easy to acquire | 0.0568 | **0.5282** | **0.2830** | **0.2921** | -0.2254 |
| Eggs_Easy to prepare | -0.0198 | -0.0551 | **0.9361** | -0.0440 | 0.0243 |
| Eggs_Family enjoys eating | **0.6204** | -0.0196 | -0.0122 | -0.0116 | -0.0227 |
| Eggs_Safe to eat | -0.0157 | 0.0354 | 0.0206 | 0.0544 | **0.9389** |
| Eggs_Is nutritious | **0.4279** | 0.0344 | 0.1368 | -0.0991 | 0.2200 |

Note: The significant domains are based on a cutoff of 0.3 for the factor loadings.

**Table S6:** PCA estimates for indicators for “Green leafy vegetables”.

| Green leafy vegetables |  |  |  |  |  |
| --- | --- | --- | --- | --- | --- |
| Component | PC1 | PC2 | PC3 | PC4 | PC5 |
| Domain | Food safety-affordability | Desirability | Accessibility | Convenience | Food quality |
| Eigenvalue | 1.78 | 1.25 | 1.16 | 0.89 | 0.81 |
| Variation explained, % | 18 | 15 | 14 | 13 | 13 |
| Indicator names |  |  |  |  |  |
| GLV_ Not expensive | **0.6653** | -0.0035 | -0.0450 | 0.0767 | -0.1067 |
| GLV_ Know vendor | -0.0927 | -0.0064 | **0.8628** | -0.0858 | 0.0467 |
| GLV_ Like the taste of | -0.0667 | **0.7246** | -0.0696 | 0.1736 | -0.0880 |
| GLV_ Easy to acquire | **0.3647** | 0.0118 | **0.4766** | 0.2604 | -0.1174 |
| GLV_ Easy to prepare | -0.0175 | 0.0046 | -0.0467 | **0.9039** | 0.0638 |
| GLV_ Family enjoys eating | 0.0718 | **0.6890** | 0.0723 | -0.1947 | 0.0978 |
| GLV_ Safe to eat | **0.6370** | -0.0057 | -0.1172 | -0.1769 | 0.1614 |
| GLV_ Is nutritious | 0.0037 | -0.0018 | 0.0203 | 0.0498 | **0.9619** |

Note: The significant domains are based on a cutoff of 0.3 for the factor loadings. GLV, green leafy vegetables.

**Table S7:**
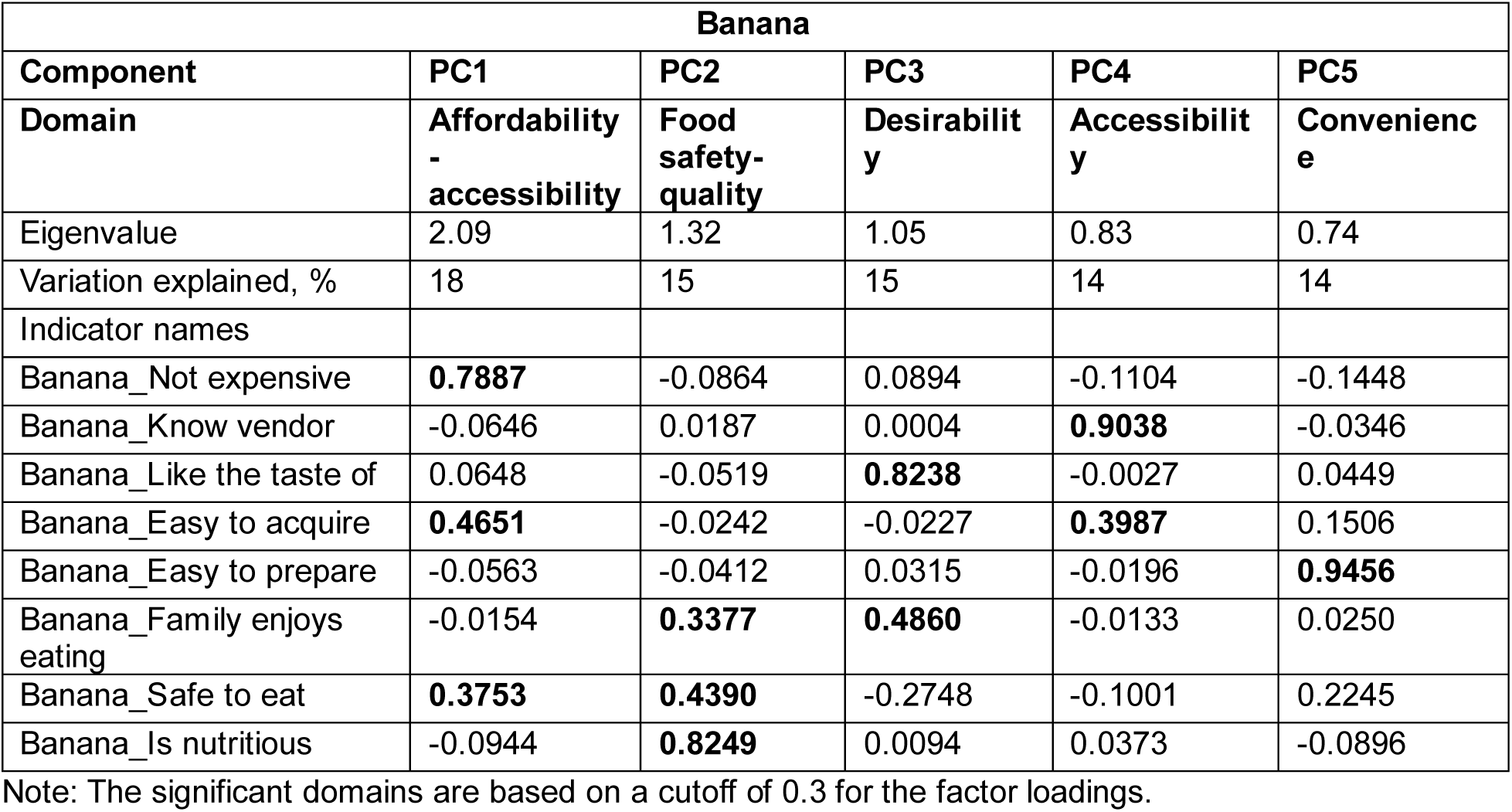
PCA estimates for indicators for “Banana”.

| Banana |  |  |  |  |  |
| --- | --- | --- | --- | --- | --- |
| Component | PC1 | PC2 | PC3 | PC4 | PC5 |
| Domain | Affordability - accessibility | Food safety-quality | Desirability | Accessibility | Convenience |
| Eigenvalue | 2.09 | 1.32 | 1.05 | 0.83 | 0.74 |
| Variation explained, % | 18 | 15 | 15 | 14 | 14 |
| Indicator names |  |  |  |  |  |
| Banana_ Not expensive | **0.7887** | -0.0864 | 0.0894 | -0.1104 | -0.1448 |
| Banana_ Know vendor | -0.0646 | 0.0187 | 0.0004 | **0.9038** | -0.0346 |
| Banana_ Like the taste of | 0.0648 | -0.0519 | **0.8238** | -0.0027 | 0.0449 |
| Banana_ Easy to acquire | **0.4651** | -0.0242 | -0.0227 | **0.3987** | 0.1506 |
| Banana_ Easy to prepare | -0.0563 | -0.0412 | 0.0315 | -0.0196 | **0.9456** |
| Banana_ Family enjoys eating | -0.0154 | **0.3377** | **0.4860** | -0.0133 | 0.0250 |
| Banana_ Safe to eat | **0.3753** | **0.4390** | -0.2748 | -0.1001 | 0.2245 |
| Banana_ Is nutritious | -0.0944 | **0.8249** | 0.0094 | 0.0373 | -0.0896 |

Note: The significant domains are based on a cutoff of 0.3 for the factor loadings.

**Table S8:** PCA estimates for indicators for “Biscuits”.

| Biscuits |  |  |  |  |  |
| --- | --- | --- | --- | --- | --- |
| Component | PC1 | PC2 | PC3 | PC4 | PC5 |
| Domain | Desirability | Convenience | Food safety-quality | Affordability | Availability |
| Eigenvalue | 2.11 | 1.30 | 1.03 | 0.86 | 0.79 |
| Variation explained, % | 21 | 14 | 14 | 13 | 13 |
| Indicator names |  |  |  |  |  |
| Biscuits_Not expensive | -0.0169 | -0.0416 | 0.0140 | **0.9671** | -0.0286 |
| Biscuits_Know vendor | -0.0189 | -0.0312 | 0.0053 | -0.0279 | **0.9716** |
| Biscuits_Like the taste of | **0.5818** | -0.1703 | 0.0388 | 0.0047 | 0.0220 |
| Biscuits_Easy to acquire | **0.3162** | **0.3464** | **-0.5666** | 0.1841 | 0.1687 |
| Biscuits_Easy to prepare | -0.0413 | **0.8869** | 0.0445 | -0.0641 | -0.0504 |
| Biscuits_Family enjoys eating | **0.6302** | 0.0227 | -0.0894 | -0.0747 | -0.1167 |
| Biscuits_Safe to eat | 0.0607 | 0.2473 | **0.6912** | 0.1405 | 0.0903 |
| Biscuits_Is nutritious | **0.3978** | -0.0110 | **0.4353** | -0.0233 | 0.0444 |

Note: The significant domains are based on a cutoff of 0.3 for the factor loadings.

**Table S9:** PCA estimates for indicators for “Fried food”.

| Fried food |  |  |  |  |  |
| --- | --- | --- | --- | --- | --- |
| Component | PC1 | PC2 | PC3 | PC4 | PC5 |
| Domain | Desirability | Food safety-quality | Accessibility | Affordability - accessibility | Convenience |
| Eigenvalue | 2.15 | 1.37 | 1.02 | 0.97 | 0.73 |
| Variation explained, % | 19 | 16 | 16 | 14 | 13 |
| Indicator names |  |  |  |  |  |
| Fried_Not expensive | 0.0200 | 0.0323 | -0.0862 | **0.8858** | -0.0140 |
| Fried_Know vendor | -0.0012 | -0.0001 | **0.8375** | -0.1577 | -0.0504 |
| Fried_Like the taste of | **0.6783** | -0.0284 | 0.0035 | 0.0449 | -0.0210 |
| Fried_Easy to acquire | 0.0114 | 0.0132 | **0.5388** | **0.3760** | 0.1083 |
| Fried_Easy to prepare | -0.0113 | -0.0177 | -0.0139 | -0.0051 | **0.9825** |
| Fried_Family enjoys eating | **0.6884** | -0.0440 | 0.0009 | 0.0040 | -0.0163 |
| Fried_Safe to eat | -0.0985 | **0.8308** | 0.0104 | 0.0920 | -0.0643 |
| Fried_Is nutritious | 0.2359 | **0.5527** | -0.0235 | -0.1965 | 0.1240 |

Note: The significant domains are based on a cutoff of 0.3 for the factor loadings.

